# A Phase III Randomized Controlled Trial Assessing the Safety and Immunogenicity of Biological E’s XBB 1.5 RBD Subunit COVID-19 Booster Vaccine in Individuals Aged 5 to 80 Years

**DOI:** 10.1101/2025.06.16.25329675

**Authors:** Subhash Thuluva, Vikram Paradkar, SubbaReddy Gunneri, Rammohan Reddy Mogulla, Vijay Yerroju, Chirag Dhar, Siddalingaiah Ningaiah, Mallikarjuna Panchakshari, Chandrashekhar S. Gillurkar, Manish Narang, Shivnitwar Sachin Kisan, A.Venkateshwar Rao

## Abstract

**Background:** The SARS-CoV-2 virus continues to evolve with recent iterations such as the Omicron sub-variants having potential for increased transmissibility. Of particular interest, the XBB.1.5 variant has been shown to cause vaccine-breakthrough cases. Biological E (BE) has utilized the same platform it used to develop CORBEVAX^TM^, an ancestral Wuhan strain COVID-19 RBD subunit vaccine (control vaccine), to now develop an XBB.1.5 RBD subunit vaccine (test vaccine).

**Methods:** To assess the safety and immunogenicity of BE’s new XBB.1.5 subunit vaccine, a prospective, randomized, single-blind Phase III study was conducted in participants aged 5 to 80 years. Participants who had not received any other approved COVID-19 vaccine within the last 6 months prior were enrolled and randomized 2:1 to receive two booster doses of either the test vaccine or the control vaccine. The vaccines were administered on Day 0 and Day 28 with immunogenicity assessments on Day 0, Day 28, and Day 42. Safety assessments included the collection of solicited and unsolicited adverse events (AEs) up until Day 56. The primary objective of the study was to show immunogenic superiority of the test vaccine booster series compared to the control vaccine series. This superiority objective was to be concluded if the lower limit of the two-sided 95% confidence interval of the anti-XBB.1.5.RBD neutralizing antibody (nAb) geometric mean titer (GMT) ratio of test:control was >1.0 on either Day 28 or Day 42. Given the emergence of JN.1 as the SARS-CoV-2 strain during the conduct of this study, Day 42 anti-JN.1 nAb were measured in a *post hoc* immunogenicity assessment. In addition, anti-XBB.1.5 RBD protein IgG concentration in the sera samples were also measured by ELISA on Day 0, Day 28, and Day 42.

**Findings:** A total of 360 participants were enrolled and randomized across 7 sites in India. The nAb GMT ratio of test:control participants was 2.08 (95% CI 1.64 to 2.63) on Day 28 and 2.91 (95% CI 2.38 to 3.56) on Day 42. The geometric mean fold rise (GMFR) of neutralizing antibodies (nAb) was 7.637 (95% CI 6.090 to 9.578) on Day 28 and 17.02 (95% CI 13.79 to 21.01) on Day 42 in the test booster series arm. The nAb GMFRs in the control booster series arm at the same time points were 3.033 (95% CI 2.340 to 3.932) and 4.824 (95% CI 3.731 to 6.236) respectively. *Post hoc* analyses revealed an nAb GMT ratio of 1.90 (95% CI 1.56 to 2.31) of test:control against the JN.1 SARS-CoV-2 strain. The safety profile of the new XBB.1.5 RBD subunit vaccine was found to be very similar to that of the Ancestral strain vaccine with 59 AEs (about 1 AE for every 8 doses administered) and 27 AEs (a little less than 1 AE for every 8 doses administered) respectively during the study. No serious AEs or AEs of special interest were reported in either the test or control arm of the study. Two cases of pyrexia were medically attended to, one in each arm.

**Interpretations:** Biological E’s new XBB.1.5 RBD subunit vaccine was found to be both safe and robustly immunogenic when administered as a two-dose booster series in 5 to 80 year olds. In particular, the vaccine induced a significant rise in neutralizing antibodies against the XBB.1.5 strain as well as cross-protective neutralizing antibodies against the JN.1 SARS-CoV-2 strain. These data are in line with studies of other XBB.1.5 monovalent vaccines and support a positive risk-benefit profile. Real world studies may provide additional evidence about the effectiveness of this new updated vaccine.

**CTRI Trial Registration Identifier:** CTRI/2024/01/061423

**Funding:** Biological E Limited.

**Research in context:** *Evidence before this study:* The SARS-CoV-2 virus has been circulating worldwide ever since its emergence in late 2019. Vaccinations have been particularly useful in preventing hospitalizations and deaths even in those that acquire COVID-19. Multiple technology platforms have been utilized to develop vaccines including mRNA and protein subunit platforms. More recently, immune-evasive SARS-CoV-2 strains have emerged fueling the need for variant-directed boosters. Multiple mRNA monovalent boosters and one protein subunit booster targeting the XBB.1.5 strain have been approved by different national regulatory agencies across the world.

*Added value of this study:* In this study, we tested a new XBB.1.5 RBD subunit vaccine that was developed using an established yeast-expression system. We ran a single-blinded randomized Phase III study to compare two booster doses of this updated vaccine versus two booster doses of the ancestral strain vaccine developed on the same yeast-expression system. We showed that the safety profile of the two vaccines was comparable. The older of these vaccines has now been safely administered to over 40,000,000 people and has been shown to have an acceptable safety profile. Additionally, the XBB.1.5 RBD subunit vaccine was found to be superior to the ancestral strain vaccine in inducing neutralizing antibodies against the XBB.1.5 strain. Importantly, this updated vaccine also induced neutralizing antibodies against the JN.1 strain, which became the most prevalent circulating SARS-CoV-2 strain during and after the conduct of the study. This study demonstrates a favourable risk-benefit profile of the updated monovalent XBB.1.5 RBD subunit vaccine when administered as a booster series across multiple age groups including children as young as 5 years.

*Implications of all the available evidence:* Our study adds to the growing body of evidence suggesting the use of updated monovalent booster vaccines will be useful in curtailing the morbidity and mortality of the SARS-CoV-2 virus. Various real-world studies have demonstrated the effectiveness of such updated vaccines in preventing hospitalizations and deaths. Protein subunit vaccines such as ours have been extensively used over the past many decades and are more likely to be accepted in an environment of growing vaccine-hesitancy. SARS-CoV-2 vaccines developed on newer platforms such as the mRNA platform and the adenoviral vector platform, while faster to market, have been shown to induce rare but serious adverse events such as myocarditis and thrombosis. Protein subunit SARS-CoV-2 vaccines do not seem to produce as many adverse events suggesting a superior safety profile.

## Introduction

About five years into the COVID-19 pandemic, the SARS-CoV-2 virus continues to evolve and presents ongoing challenges to global public health efforts. While COVID-19 has transitioned towards an endemic status, the emergence of novel Omicron sub variants, particularly the XBB lineages, has driven new surges of infections worldwide ^1, 2^. The Omicron variant (B.1.1.529) had been the predominant SARS-CoV-2 strain in circulation since January 2022, spawning over 680 sub-lineages ^3^. Its high transmissibility and capacity for immune evasion had resulted in widespread vaccine-breakthrough infections and reinfections ^4, 5^. Among the Omicron subvariants, the World Health Organization (WHO) designated BQ.1.1 and XBB* (XBB and its sub-lineages, including XBB.1.5) as “Omicron subvariants under monitoring” due to their potential for increased transmissibility and immune escape ^6^. Fortunately, no significant increase in disease severity has been observed to date despite these evolving variants.

Of particular concern is the XBB.1.5 subvariant, a recombinant of two Omicron predecessors, BA.2.10.1 and BA.2.75. Preliminary research indicates that XBB.1.5 exhibits enhanced immune evasion properties and increased binding affinity to the human ACE2 receptor compared to other circulating Omicron subvariants ^7, 8^. The F486P mutation in the receptor-binding domain of the spike glycoprotein distinguishes XBB.1.5 from its XBB or XBB.1 predecessor and has been associated with increased ACE2 binding ^9^. XBB was the most prevalent sub-lineage (63.2 per cent) circulating all over India as of December 2023. The World Health Organization (WHO) TAG-CO-VAC recommended that new formulations of COVID-19 vaccines should aim to induce antibody responses that neutralize XBB descendent lineages in order to improve protection against XBB.1-related variants. One approach recommended was to use a monovalent XBB.1 descendent lineage, such as XBB.1.5 as the vaccine antigen ^10^.

Biological E (BE) presently has a subunit COVID vaccine, CORBEVAX^TM^ (targeting the ancestral Wuhan strain) that has market authorization in India and has been granted Emergency Use Listing by the WHO ^11, 12, 13^.

To contain the spread of the new variant, BE developed a new XBB.1.5-RBD subunit vaccine on the same platform used for the vaccine against the ancestral strain. Animal toxicity and immunology studies indicated a favourable risk-to-benefit profile (unpublished data) and the vaccine was advanced for clinical development. This manuscript describes the design and results of a Phase III safety and immunogenicity study of BE’s XBB.1.5-RBD vaccine in individuals aged ≥5 to ≤80 years, when administered as a booster series.

## Methods

### Trial Design and Setting

This study was a prospective, multicenter, single-blind, randomized, comparative phase III clinical trial conducted at 7 study sites across India between January 2024 and June 2024 (site names and locations provided in the supplementary data). The study adhered to the principles outlined in the Declaration of Helsinki, the International Conference on Harmonisation guidelines for Good Clinical Practices (GCP), and local regulatory standards. The protocol received approval from the Investigational Review Board or Ethics Committee at each study site. All participants and/or their parents gave their written informed consent prior to enrollment. No patients or members of the public were involved in the design, conduct, reporting, or dissemination of this research. All study sites were required to have the capacity to store and administer investigational vaccines under cold chain conditions and staff qualified in GCP-compliant clinical trial procedures. Vaccinations were administered by trained personnel licensed according to local regulatory requirements.

The total duration of the study for each participant was 56 days from the day of the first dose of vaccination. Study participants received two doses of test or control vaccine, administered on Day 0 and Day 28. Three participants did not receive the second dose of the vaccine as they dropped out of the study before Day 28. Immunogenicity was assessed at Day 0 (pre-vaccination), Day 28 and at Day 42 (14 days post the second dose) while the safety profile was assessed until 28 days after the second dose (Day 56). A time window of +4 days was allowed for Day 28 and Day 42 visits and a time window of 14 days was allowed for the Day 56 visit to ensure participant compliance.

During the course of this study, no major protocol deviations were reported at any of the study sites. A few participants reported for their visits out of the window period but these deviations were not found to be significant and were duly reported to the ethics committees of the respective study sites. No formal interim efficacy or safety analyses or predefined stopping rules were included in the protocol.

### Participants

A total of 360 healthy individuals, aged between 5 and 80 years were enrolled in the study. The participants were further stratified into three age subsets: ≥50 to ≤80 years (older adults), ≥18 to ≤49 years (younger adults) and ≥5 to ≤17 years (children/adolescents). Within each age subset, participants were randomized in a 2:1 ratio between test and control vaccine groups.

Medically stable individuals who were previously vaccinated with any existing COVID-19 vaccine – either as a primary series only or a primary series and a booster dose, with the most recent dose at least six months prior to enrolment – were eligible to participate in the study. Health status was assessed during the screening period and included an assessment of medical history, clinical laboratory tests, vital signs, and a physical examination. Other key eligibility criteria included being virologically negative for SARS-CoV-2 infection confirmed by an RT-PCR test at the screening visit (day -1 to -3), seronegative to HIV 1 & 2, HBV and HCV infection. All those with an axillary temperature of more than 38·0°C, those participating in any other clinical trial, those with a known allergy to vaccine components, those living in the same household of SARS-CoV-2 positive person(s), those with symptoms suggestive of an acute illness, and those with other chronic illnesses/immunodeficient conditions or on immunosuppressants were excluded from the study. A complete list of eligibility criteria is provided as supplementary information. Licensed or investigational COVID-19 vaccines were prohibited during study follow-up. Medications for symptom relief were permitted, but immunoglobulins or blood products were not allowed within the 3 months prior to enrollment.

### Randomization, Allocation Concealment, and Blinding

Enrolled participants were randomized in a 2:1 ratio to either receive the XBB.1.5 RBD subunit vaccine (test) or CORBEVAX^TM^ ancestral strain RBD vaccine (control). An interactive web response system-based (IWRS) platform was utilized for randomization, that occurred after completion of all screening-related activities and prior to the administration of first booster dose. A participant was considered randomized once they met all the eligibility criteria and received a randomization number from the IWRS platform. The randomization scheme was generated by SAS statistical program. Site staff accessed treatment assignments through the IWRS interface, and allocation was concealed until the point of assignment. This was a single-blind study where study participants were unaware of the vaccination group to which they were assigned. The test and control vaccines were identical in appearance, packaging, and labeling to ensure blinding of participants.

### Interventions (Vaccines)

Biological E’s CORBEVAX^TM^ – a subunit vaccine containing the receptor binding domain (RBD) of the ancestral SARS-CoV-2 strain – was used as a control vaccine. As a test vaccine, a new formulation was developed that had the XBB.1.5 RBD antigen of SARS-CoV-2. Both RBD proteins were produced on the same *Pichia pastoris* yeast platform and both the formulated vaccines contained the RBD protein antigen and Aluminum Hydroxide gel as Al^3+^ and CpG1018 as adjuvants ^14^. A 0·5 mL dose of the candidate XBB.1.5 RBD antigen of SARS-CoV-2 test vaccine or the CORBEVAX^TM^ vaccine containing RBD from the ancestral strain (control vaccine) was administered via an intramuscular (IM) injection into the deltoid muscle of the non-dominant arm in a two-dose booster schedule with a 28 day interval between doses. Both the vaccines contained 25 µg of RBD Antigen, 750 µg of Aluminium Hydroxide (as Al^3+^) and 750 µg of CpG1018.

### Outcomes

This Phase III study was designed to assess both the immunogenicity and safety of BE’s new XBB.1.5 subunit COVID-19 test vaccine. The primary objective was to assess the immunogenic superiority against the monovalent CORBEVAX^TM^ control vaccine (containing the ancestral Wuhan strain) administered in two doses, with 28 days interval between doses, in terms of virus neutralizing antibodies at Day 28 and Day 42. The primary endpoint was the geometric mean titer (GMT) of anti-XBB.1.5.RBD neutralizing antibodies at Day 28 and at Day 42 in cohorts that received the test and control vaccine for superiority demonstration. Superiority was to be concluded if the lower limit of the two-sided 95% confidence interval of the GMT ratio of test:control of anti-XBB.1.5.RBD neutralizing antibody titers was >1.0 either at Day 28 or Day 42. The secondary endpoint was to assess GMT at baseline, and to calculate the geometric mean fold rise (GMFR) at Day 28 and at Day 42 in both the test and control group. Another secondary immunogenicity endpoint was to calculate the proportion of participants at Day 28 and Day 42 with a ≥2-fold rise of neutralizing antibodies in those with pre-existing antibody titers, and a ≥4-fold rise in those without pre-existing antibody titers when compared with baseline titers.

The secondary objective was to descriptively assess the overall safety, reactogenicity and tolerability of BE’s XBB.1.5-RBD subunit Covid-19 test vaccine in comparison with CORBEVAX^TM^ control vaccine (containing the ancestral Wuhan strain) for a period of 28 consecutive days after each dose in all enrolled individuals. The secondary endpoints were defined as occurrence of any adverse reactions within 60 minutes post-vaccination after each booster dose, occurrence of any solicited adverse events (AEs) within 7 consecutive days after both doses, and occurrence of any unsolicited AEs; medically attended AEs; AEs of special interest in the 28 days’ post-vaccination period.

In April 2024 – about midway through the conduct of this study - the WHO’s TAG-CO-VAC team, determined that the JN.1 strain of SARS-CoV-2 was the dominant strain circulating worldwide. Hence, additional *post hoc* outcomes relating to protection against the JN.1 strain were included in this study. Briefly, neutralizing antibodies directed against the JN.1 strain were assessed on Day 0, Day 28, and Day 42 in the test and control groups, and descriptively compared.

### Safety Assessments

The safety assessments were based on a descriptive comparison of incidence rates of solicited local and systemic AEs up to 7 days (Day 0-6), captured through subject diary after each vaccine dose. Participants were observed for 60 minutes post-vaccination to assess reactogenicity. Solicited local AEs were pain, redness, swelling, itching or warmth at injection site. Solicited systemic AEs were fever, headache, chills, Myalgia, arthralgia, fatigue, nausea, urticaria, rhinorrhea, irritability, hypotonic-hyporesponsive episodes, somnolence, seizure, and acute allergic reaction. Unsolicited adverse events (AEs) were assessed up to Day 28 after each dose and serious AEs (SAEs), medically attended AEs (MAAEs), and AEs of special interest (AESIs) were followed up throughout the study duration.

### Immunogenicity Assessments

#### Detection of anti-XBB.1.5 RBD IgG concentration

The concentration of IgG that specifically bound to the XBB.1.5 receptor-binding domain (RBD) protein in the sera samples was measured in a sandwich ELISA. Purified XBB.1.5 RBD protein was bound to the 96-well plate and the sera samples were added to the wells to enable attachment of the RBD specific IgGs. After washing the plate, a secondary antibody directed against Human IgG and linked to an enzyme was added to the plate. Finally, the enzyme substrate was added resulting in a reaction that produced a color signal which was detected and quantified by spectrophotometry. The intensity of the color is directly proportional to the concentration of RBD specific IgG in the sera sample. A pooled serum sample was used as a reference standard for comparison and the anti-RBD IgG concentration is reported in ELISA Units/mL (EU/mL). Day 0, Day 28 and Day 42 sera samples of all the clinical trial participants were tested for anti-RBD IgG concentration by ELISA.

#### Detection of anti-XBB.1.5 neutralizing antibody

Neutralizing antibodies (nAbs) were measured to assess level of antibodies that were capable of blocking the SARS-CoV-2-XBB.1.5 strain from infecting mammalian cells. To enable testing in conventional laboratories, a pseudovirus based on the HIV-1 Lentivirus expressing the spike protein prepared via transfection was utilized in this study ^15^. This Lentivirus expressed the Spike Protein from the SARS-CoV-2-XBB.1.5 strain as well as a reporter enzyme viz. Nano-luciferase. This XBB.1.5-Pseudovirus (PSV) was shown to infect HEK-293T-ACE2 cells, a human cell line that overexpresses the ACE-2 receptor. Once the pseudovirus infects cells, the enzyme Nano-luciferase is produced and can convert the specific substrate into a reaction product that can be detected by a chemiluminescence detector. When the PSV were pre-incubated with sera samples, the nAbs present in the sera blocked the PSV’s ability to infect the HEK-293T-ACE2 cells thereby reducing the chemiluminescence signal directly proportional to the degree of neutralization. Thus, by conducting neutralization of the PSV at different dilutions of the sera and measuring the observed PSV neutralization, a dilution (i.e. titer) of the sera that represented 50% neutralization of the PSV added to the cells was calculated and termed as Pseudo-Virus Neutralization Titer50 or PSVNT (directly proportional to the nAb titer in the sera sample). Day 0, Day 28 and Day 42 sera samples from all the clinical trial participants were tested for PSVNT.

#### Detection of anti-JN.1 neutralizing antibodies and anti-JN.1 RBD IgG antibodies

The JN.1-PSV consisted of the same HIV-1 Lentivirus that was utilized to generate the PSV representing the XBB.1.5 strain, except for the use of the JN.1 spike protein sequence. Day 0 and Day 42 sera samples collected from the study were tested and nAb titers against the JN.1-PSV were determined. Similar statistical analysis, as performed for XBB.1.5-PSV nAb titers, was also performed for JN.1-PSV nAb titers.

#### Sample Size calculations

Sample size and power were estimated to evaluate superiority of the XBB.1.5-RBD vaccine over the control using a one-sided, two-sample t-test on log-transformed neutralizing antibody titers under a parallel-group design. The primary hypothesis tested whether the GMT ratio (test/control) exceeded 1.0, with superiority concluded if the lower bound of the two-sided 95% confidence interval was >1.0. The calculation assumed a log-transformed mean difference of 0.5 (corresponding to a GMT ratio of approximately 1.64), selected *a priori* to reflect a realistic and clinically meaningful difference in neutralizing antibody levels between the test and control vaccines. A geometric standard deviation of 3.9 (log SD ≈ 1.36), derived from prior in-house Omicron-specific immunogenicity data, and a 2:1 randomization ratio were used. A total of 240 participants in the test group and 120 in the control group (inclusive of 10% dropout) provided 90.005% power at a one-sided 2.5% significance level. The sample size was finalized after trial-and-error converged on a total N of 350. Assuming actual dropout remained below 10 participants, but accounting for a 10% contingency (i.e. 36 participants), the initial design assumption was subsequently back-calculated against N = 360, yielding an implied log-transformed effect size of 0.493, corresponding to a GMT ratio of 1.637.

### Statistical Methods

Three populations were pre-specified for statistical analyses: the intention-to-treat (ITT) population (all randomized participants), the safety population (those who received at least one vaccine dose), and the according-to-protocol (ATP) population (participants who completed both doses, had no major protocol deviations, and contributed valid immunogenicity data at all protocol-defined timepoints).

Primary immunogenicity analyses for demonstration of superiority were conducted using the ATP population. Descriptive analyses (e.g., demographics, baseline characteristics) were performed using the ITT population while safety analyses were performed on the safety population. Safety analyses included descriptive summaries of solicited and unsolicited adverse events, serious adverse events, and adverse events of special interest. Missing safety and immunogenicity data were not imputed; all analyses were based on observed and valid data only.

For immunogenicity analyses, the geometric mean titers (GMT) of the virus neutralization antibodies for each of the age groups was calculated separately. A two-sided 95% confidence interval (CI) for the post vaccination geometric mean titers was calculated. Immunogenicity was compared using a two-sample t-test on the means of log-transformed titers at the 2.5% significance level (one-sided) in order to allow for pairwise comparisons. For each comparison, the ratio of the GMTs along with their corresponding 95% CI was presented. There was no formal allowance for multiplicity in testing two primary outcomes (Day 28 and Day 42 GMTs). All the immunogenicity data were log transformed and are anti-logged before interpretation.

### Ethical Considerations

The study was conducted in accordance with the principles defined in the Declaration of Helsinki, International Conference on Harmonization guidelines (Good Clinical Practices), and the New Drugs and Clinical Trial Rules, 2019 from the Central Drugs Standard Control Organization of India. The investigational review board or ethics committee at each study site approved the protocol.

## Results

### Study Population and Disposition

A total of 372 children and adults were screened of whom 360 participants were enrolled with 240 randomized to the test vaccine and 120 to the control vaccine. The mean age in sub group-1, -2, and -3 was 55.41 years, 32.53 years, 13.01 years in the group that received the test vaccine, and 55.78 years, 34.06 years, and 13.35 years in the group that received the control vaccine. All participants were Indian by nationality. Participants were predominantly male, comprising nearly 68% in the test vaccine group and 65% in the control vaccine group. These data are summarized in Table 1 and Supplementary Table 1. All 360 participants had received 2 doses of a primary COVID-19 vaccine series and about 1 in 6 participants had received a booster dose as well.

**Table 1:**
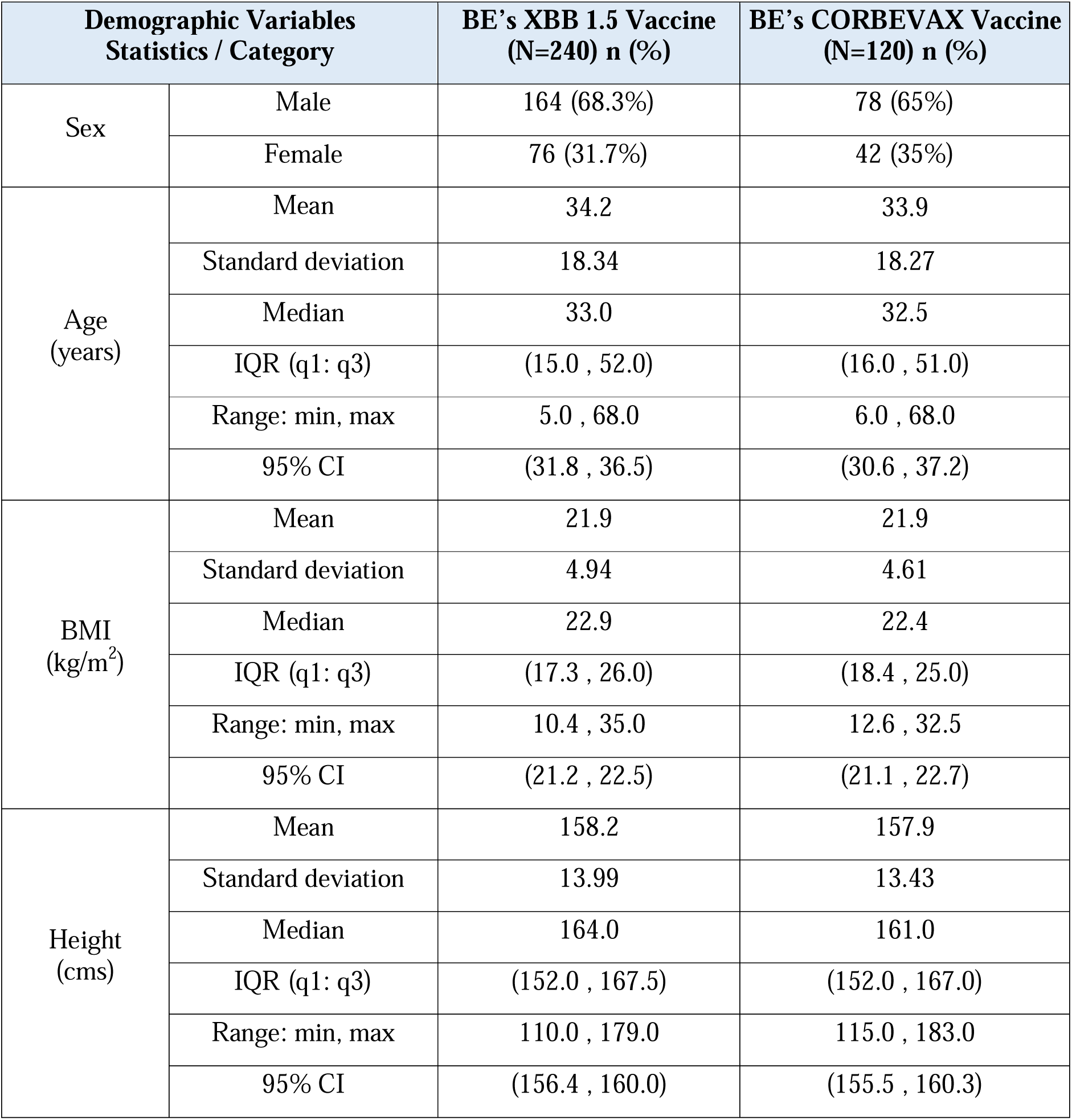
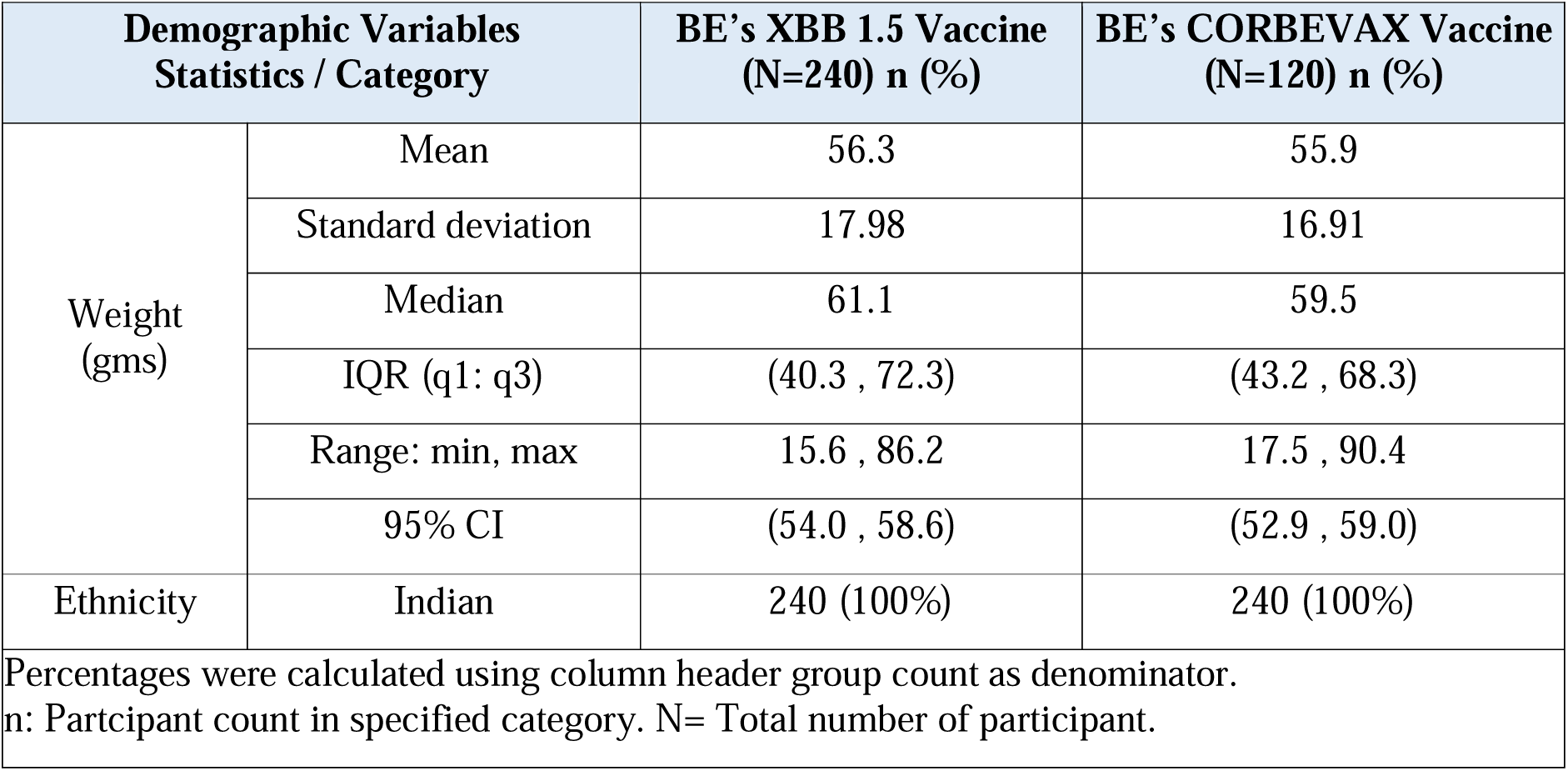
Demographic characteristics.

| Demographic Variables<br>Statistics / Category |  | BE's XBB 1.5 Vaccine<br>(N=240) n (%) | BE's CORBEVAX Vaccine<br>(N=120) n (%) |
| --- | --- | --- | --- |
| Sex | Male | 164 (68.3%) | 78 (65%) |
|  | Female | 76 (31.7%) | 42 (35%) |
| Age<br>(years) | Mean | 34.2 | 33.9 |
|  | Standard deviation | 18.34 | 18.27 |
|  | Median | 33.0 | 32.5 |
|  | IQR (q1: q3) | (15.0 , 52.0) | (16.0 , 51.0) |
|  | Range: min, max | 5.0 , 68.0 | 6.0 , 68.0 |
|  | 95% CI | (31.8 , 36.5) | (30.6 , 37.2) |
| BMI<br>(kg/m<sup>2</sup>) | Mean | 21.9 | 21.9 |
|  | Standard deviation | 4.94 | 4.61 |
|  | Median | 22.9 | 22.4 |
|  | IQR (q1: q3) | (17.3 , 26.0) | (18.4 , 25.0) |
|  | Range: min, max | 10.4 , 35.0 | 12.6 , 32.5 |
|  | 95% CI | (21.2 , 22.5) | (21.1 , 22.7) |
| Height<br>(cms) | Mean | 158.2 | 157.9 |
|  | Standard deviation | 13.99 | 13.43 |
|  | Median | 164.0 | 161.0 |
|  | IQR (q1: q3) | (152.0 , 167.5) | (152.0 , 167.0) |
|  | Range: min, max | 110.0 , 179.0 | 115.0 , 183.0 |
|  | 95% CI | (156.4 , 160.0) | (155.5 , 160.3) |
| Weight<br>(gms) | Mean | 56.3 | 55.9 |
|  | Standard deviation | 17.98 | 16.91 |
|  | Median | 61.1 | 59.5 |
|  | IQR (q1: q3) | (40.3 , 72.3) | (43.2 , 68.3) |
|  | Range: min, max | 15.6 , 86.2 | 17.5 , 90.4 |
|  | 95% CI | (54.0 , 58.6) | (52.9 , 59.0) |
| Ethnicity | Indian | 240 (100%) | 240 (100%) |
| Percentages were calculated using column header group count as denominator.<br>n: Participant count in specified category. N= Total number of participant. |  |  |  |

Of the 240 that received the test vaccine, 1 participant was lost to follow-up. Similarly, 2 of the 120 that received the control vaccine were lost to follow up. Therefore, all immunogenicity analyses on Day 28 and Day 42 included 357 participants while Day 0 included all 360 participants (CONSORT flowchart of study participants shown in Figure 1).

**Figure 1:**
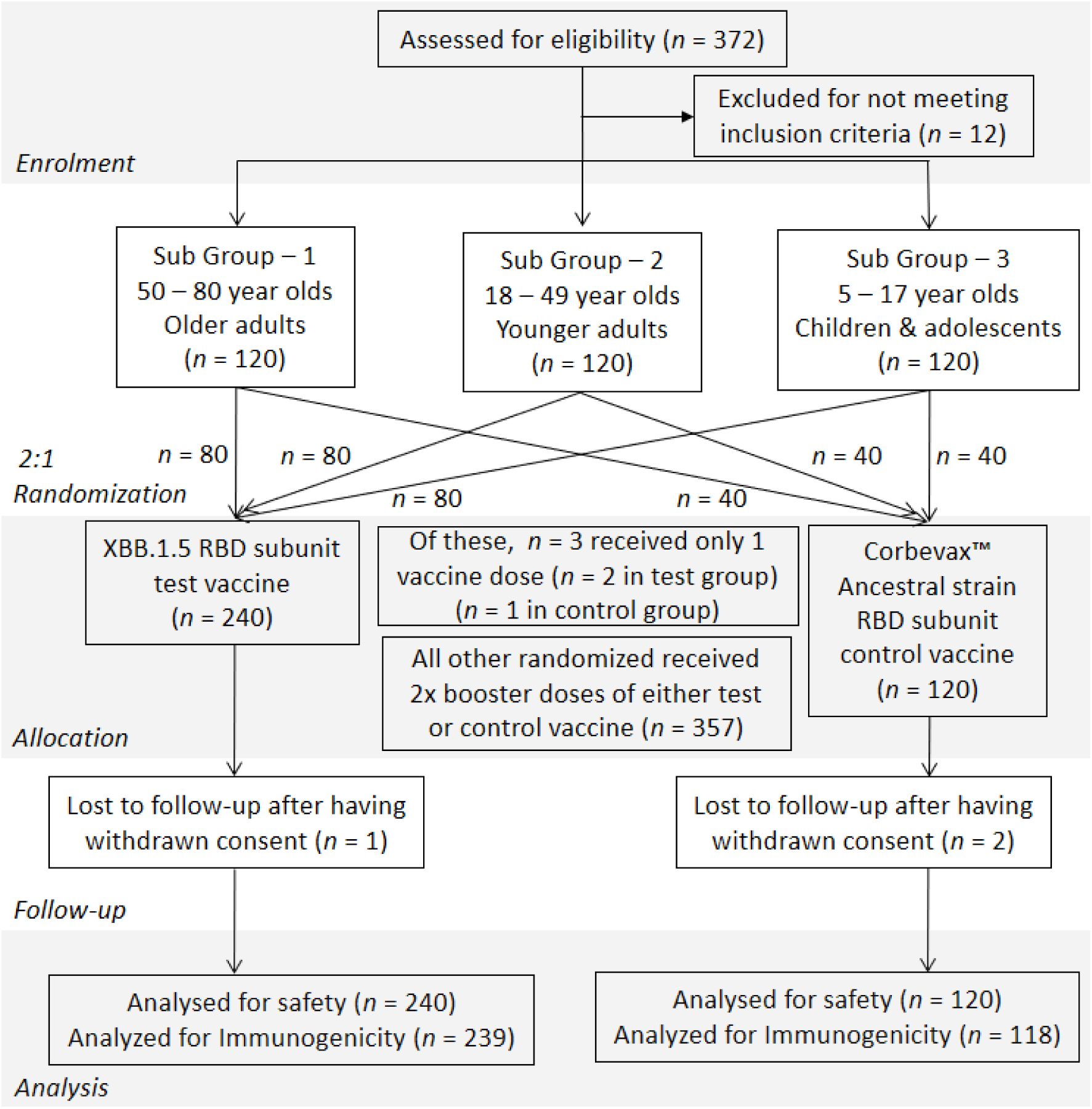
CONSORT 2025 study flowchart.

### Immunogenicity findings

#### XBB.1.5 subunit test vaccine induced superior immune responses

The primary objective of this Phase III study was to assess the immunogenicity of the XBB.1.5 subunit test vaccine in comparison with Corbevax^TM^ control vaccine directed against the ancestral COVID-19 strain. As primary endpoint, the neutralizing antibody titers against the XBB.1.5 strain were measured on Day 28 and Day 42 in participants that either received the test or control vaccine (Figure 2a). Superiority was to be determined and concluded if the lower limit of the 95% confidence interval of the test : control GMT was >1 on either Day 28 or Day 42. As shown in Table 2, this ratio was 2.08 (lower limit of 1.64) on Day 28 and 2.91 (lower limit of 2.38) on Day 42 (Figure 2b). These data show that the updated XBB.1.5 subunit vaccine is superior to the vaccine directed against the ancestral Wuhan strain.

**Figure 2:**
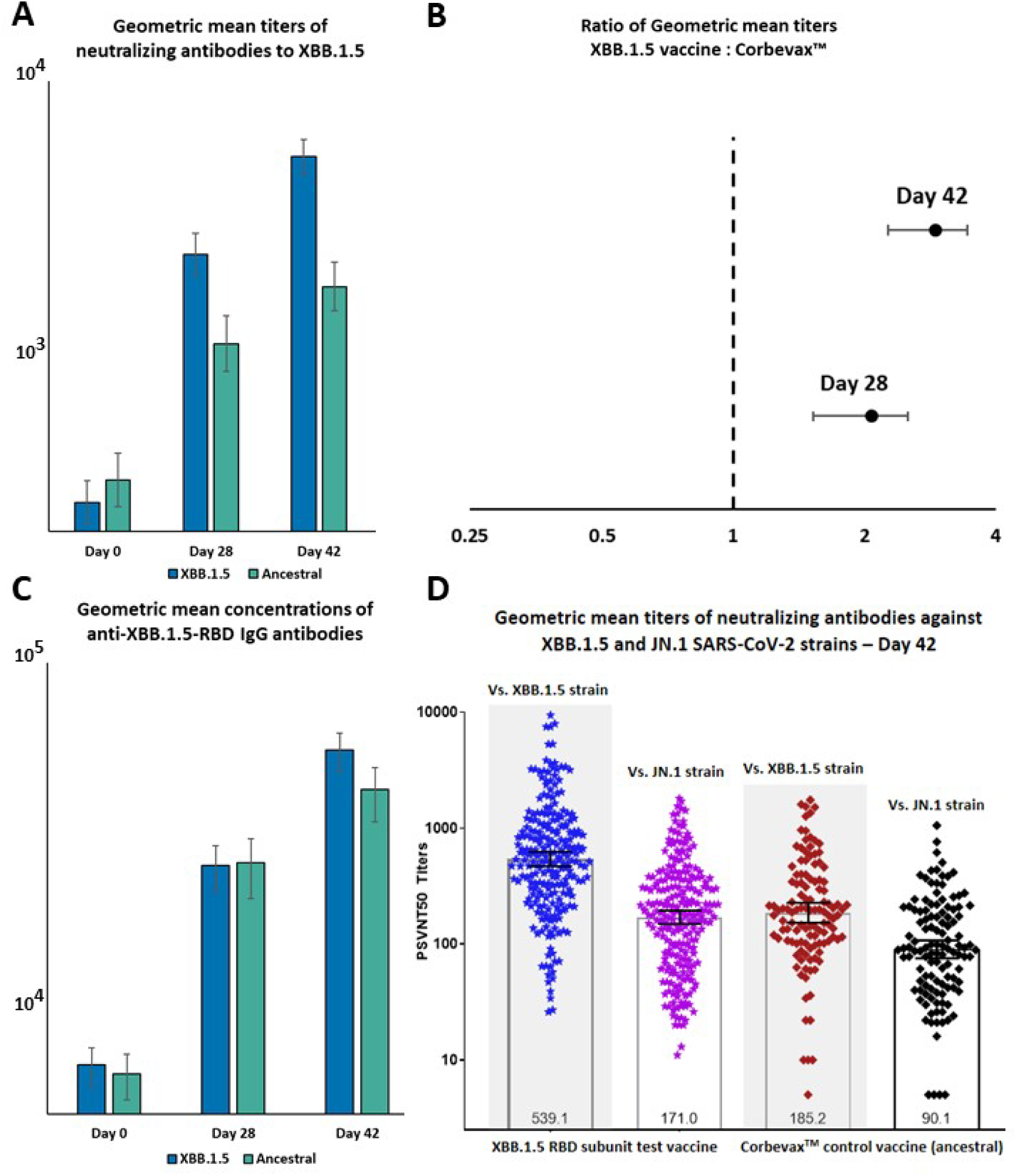
A) Geometric mean titers B) Geometric mean titer ratio of XBB.1.5 neutralizing antibodies. C) Geometric mean concentration anti-XBB.1.5 IgG antibodies (“XBB.1.5” represents the XBB.1.5 RBD subunit test vaccine and “Ancestral” represents CORBEVAX^TM,^ the Ancestral SARS-CoV-2 strain RBD subunit control vaccine) D) Geometric mean titers of neutralizing antibodies measured by PSVNT 50 induced against the JN.1 and XBB.1.5 strains of SARS-CoV-2 14 days after the two-dose booster series.

**Table 2:**
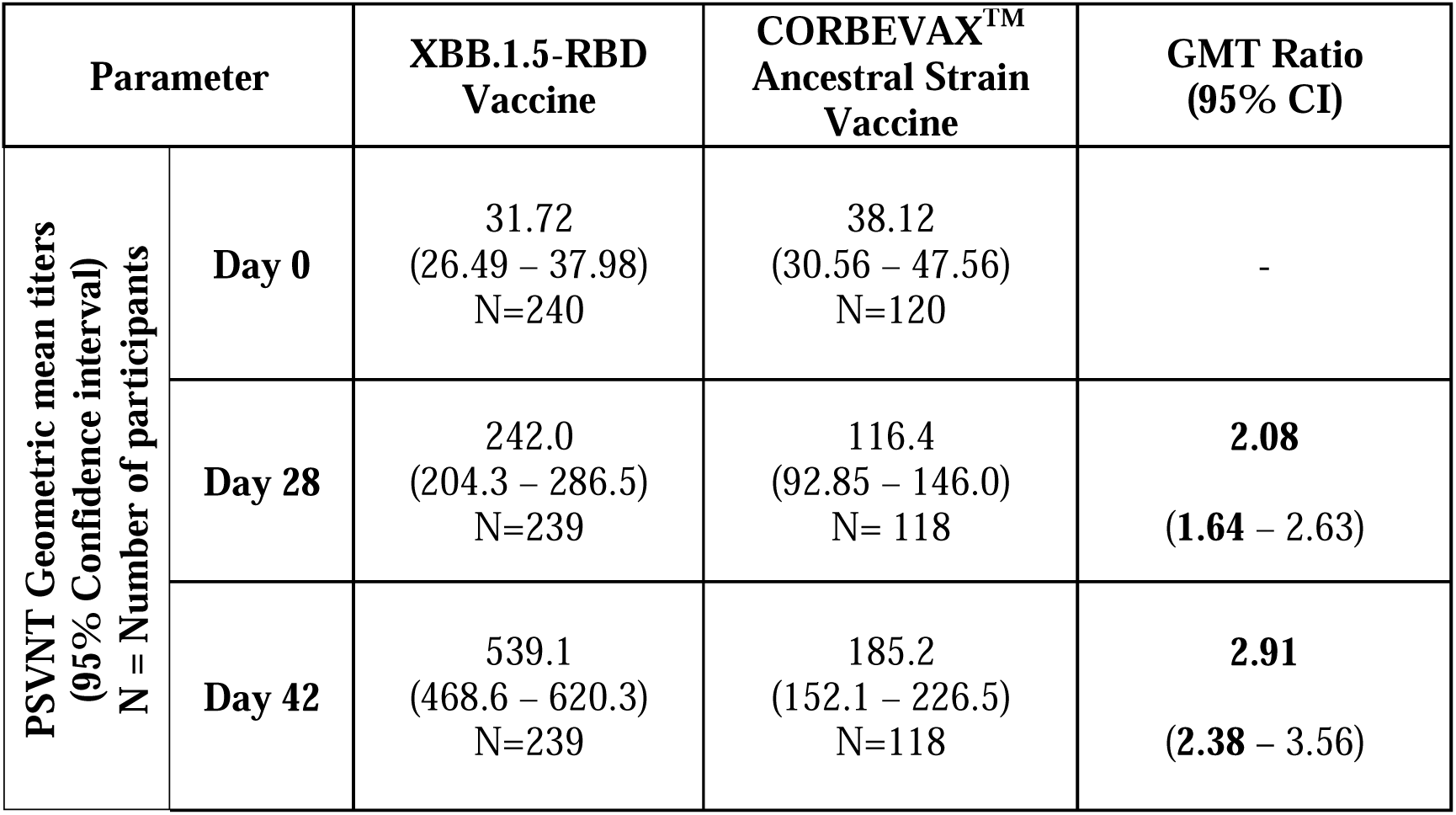
Anti-XBB.1.5 geometric mean neutralizing antibody titer (GMT) ratios of XBB.1.5 arm : CORBEVAX^TM^ at different time points.

One of the secondary immunogenicity objectives was to assess the GMFR of both nAbs as well as IgGs. These data are presented in Table 3 and show that the GMFR for neutralizing antibodies on Day 28 is nearly 8 after the XBB.1.5 subunit vaccine while only about 3 after CORBEVAX^TM^. At Day 42, the GMFR levels are more than 17 after the XBB.1.5 subunit vaccine and about 5 after CORBEVAX^TM^. The GMFR differences were more modest when it came to IgG antibody levels with a GMFR of about 8 on Day 42 after the XBB.1.5 subunit vaccine while a little more than 6.5 after CORBEVAX^TM^. (Geometric mean concentrations of anti-XBB.1.5-RBD IgGs presented in Figure 1c).

**Table 3:** Geometric mean fold rise (GMFR) of neutralizing antibody and anti-XBB.1.5 IgG titers in the two arms at different time points.

| GMFR (95% CI) |  | XBB.1.5 subunit test vaccine | CORBEVAX™ control vaccine |
| --- | --- | --- | --- |
| Neutralizing antibodies (PSVNT) | Day 28 | 7.637 (6.090 – 9.578) | 3.033 (2.340 – 3.932) |
|  | Day 42 | **17.02** (13.79 – 21.01) | **4.824** (3.731 – 6.236) |
| Anti-XBB.1.5 IgG antibodies by ELISA | Day 28 | 3.772 (3.208 – 4.434) | 4.047 (3.284 – 4.986) |
|  | Day 42 | 8.130 (6.975 – 9.476) | 6.575 (5.356 – 8.070) |

Lastly, the proportion of participants that underwent seroconversion on Day 28 (after dose -1) and on Day 42 (after dose -2) were assessed. For the XBB.1.5 subunit vaccine, 79.1% and 89.1% were seroconverted by nAbs on Day 28 and Day 42 respectively. For the Corbevax^TM^ arm, the same seroconversion rates 58.5% and 68.6% respectively. Anti-XBB.1.5 IgG seroconversion rates were comparable in the two arms at both time points. Additional measures of 2- and 4-fold-rise in neutralizing antibodies are provided in Table 4.

**Table 4:**
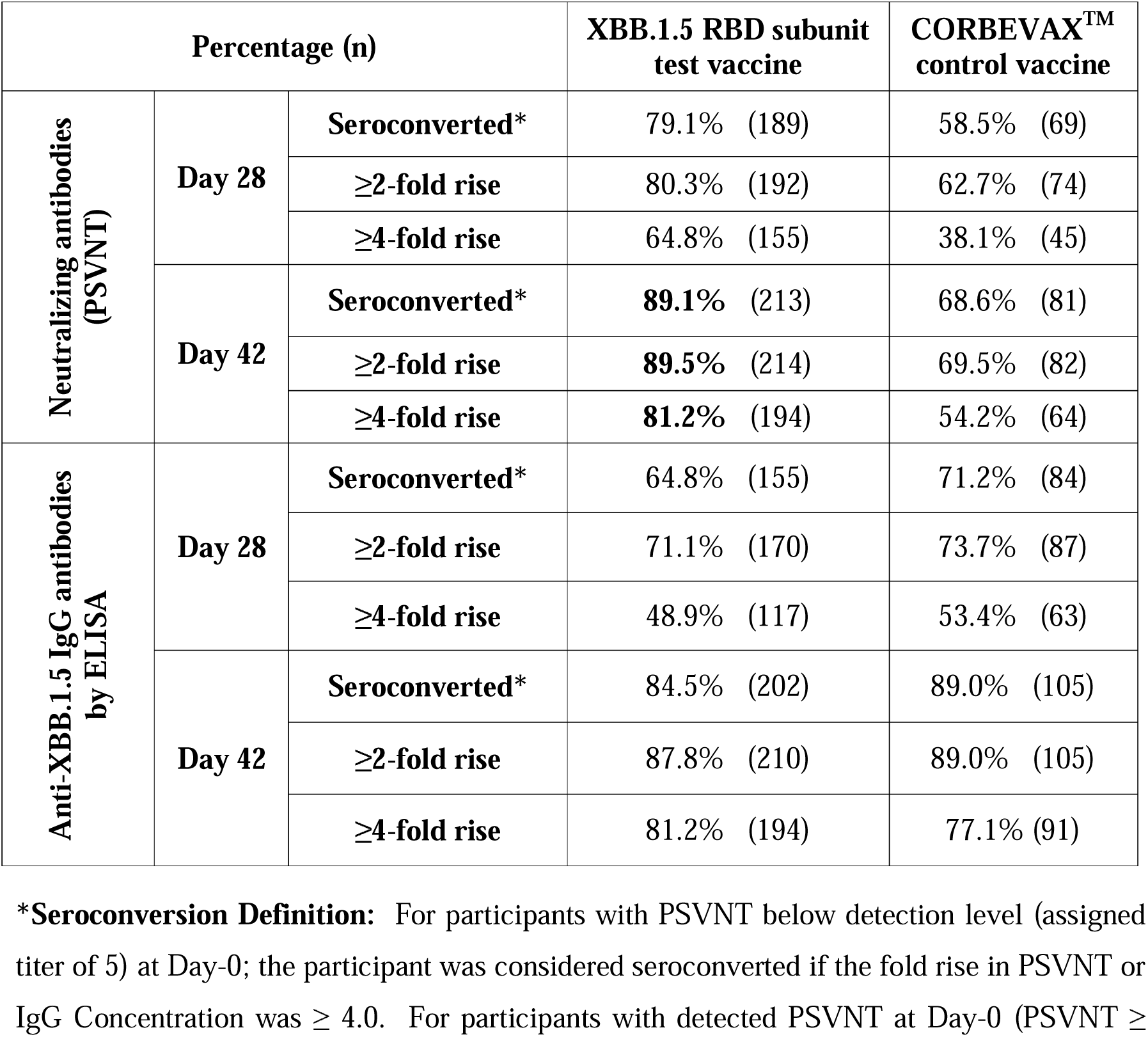
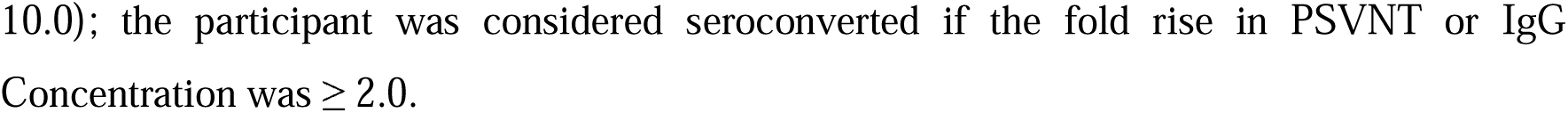
Percentage of participants that were seroprotected, with a≥2-fold rise, and ≥4-fold rise in neutralizing antibodies after the test and control vaccine on Day 28 and Day 42.

| Percentage (n) |  |  | XBB.1.5 RBD subunit test vaccine | CORBEVAX™ control vaccine |
| --- | --- | --- | --- | --- |
| Neutralizing antibodies (PSVNT) | Day 28 | Seroconverted* | 79.1% (189) | 58.5% (69) |
| | | $\geq 2$-fold rise | 80.3% (192) | 62.7% (74) |
| | | $\geq 4$-fold rise | 64.8% (155) | 38.1% (45) |
|  | Day 42 | Seroconverted* | **89.1%** (213) | 68.6% (81) |
| | | $\geq 2$-fold rise | **89.5%** (214) | 69.5% (82) |
| | | $\geq 4$-fold rise | **81.2%** (194) | 54.2% (64) |
| Anti-XBB.1.5 IgG antibodies by ELISA | Day 28 | Seroconverted* | 64.8% (155) | 71.2% (84) |
| | | $\geq 2$-fold rise | 71.1% (170) | 73.7% (87) |
| | | $\geq 4$-fold rise | 48.9% (117) | 53.4% (63) |
|  | Day 42 | Seroconverted* | 84.5% (202) | 89.0% (105) |
| | | $\geq 2$-fold rise | 87.8% (210) | 89.0% (105) |
| | | $\geq 4$-fold rise | 81.2% (194) | 77.1% (91) |

**\*Seroconversion Definition:** For participants with PSVNT below detection level (assigned titer of 5) at Day-0; the participant was considered seroconverted if the fold rise in PSVNT or IgG Concentration was $\geq 4.0$ . For participants with detected PSVNT at Day-0 (PSVNT ≥
10.0); the participant was considered seroconverted if the fold rise in PSVNT or IgG Concentration was $\geq 2.0$ .

#### XBB.1.5 subunit vaccine induced cross-reactive neutralizing antibodies against the JN.1 strain

Given the emergence of JN.1 during the conduct of the study, neutralizing antibodies titers against the JN.1 strain were measured on Day 0 and Day 42 in both arms of the study. *Post hoc* analyses of these data showed that the XBB.1.5 RBD subunit vaccine induced nearly 2-times the levels of neutralizing antibodies against the JN.1 strain than the ancestral strain control vaccine (GMT ratio of 1.90 with 95% CI from 1.56 to 2.31). This finding meets the *a priori* superiority criteria drawn up for the XBB.1.5 strain, suggesting significant cross-neutralization against the JN.1 strain as well (Table 5).

**Table 5:** Anti-JN.1 geometric mean neutralizing antibody titer (GMT) ratio of XBB.1.5 arm : CORBEVAX^TM^ at Day 42 (*Post hoc* analysis)

| Parameter | XBB.1.5-RBD Vaccine | CORBEVAX™ Ancestral Strain Vaccine | GMT Ratio (95% CI) |
| --- | --- | --- | --- |
| **Day 42 PSVNT against JN.1** | 171.0<br>(149.2 – 196.0)<br>N=239 | 90.1<br>(74.44 – 109.0)<br>N=118 | **1.90**<br>**(1.56 – 2.31)** |

The individual titers and geometric mean titers of those vaccinated with either the XBB.1.5 RBD subunit vaccine or control vaccine against both the XBB.1.5 and JN.1 stains is presented in Figure 2D. The GMTs induced by the XBB.1.5 RBD subunit vaccine against the JN.1 strain were significantly higher than those induced by the ancestral strain vaccine.

These immunogenicity data when taken together point towards enhanced neutralization ability by the XBB.1.5 subunit vaccine compared to the CORBEVAX^TM^ control ancestral Wuhan strain vaccine. Additionally, the XBB.1.5 RBD subunit vaccine also provides significant neutralization of the JN.1 strain of SARS-CoV-2 as well.

### Safety findings

Overall, 59 adverse events (AEs) were reported after the XBB.1.5-RBD subunit vaccine with these events being reported in 54 (22.50%) participants. The most frequently reported AEs were injection site pain [30 events in 27 (11.25%) participants], pyrexia [13 events in 13 (5.42%) participants], and headache [9 events in 9 (3.75%) participants]. The unsolicited AEs reported after the XBB.1.5-RBD subunit vaccine were headache [2 events in 2 (0.83%) participants], and pyrexia, fatigue, nausea, abdominal pain, cough and rhinorrhea all reported in 1 participant each [1 event in 1 (0.41%) participant each].

On the other hand, 27 AEs were reported after CORBEVAX^TM^, the ancestral SARS-CoV-2 strain control vaccine in 27 (22.50%) participants. The most frequently reported AEs in this control arm were injection site pain [14 events in 14 (11.67%) participants], pyrexia [5 events in 5 (4.16%) participants], injection site erythema, and headache [3 events in 3 (2.50%) participants each]. Unsolicited AEs reported after CORBEVAX^TM^ were miliaria (prickly heat), headache, fatigue, pyrexia, and injection site pain [1 event in 1 (0.83%) participant each].

Importantly, no serious AEs were reported in either arm of the study. Only 2 AEs required medical attention of any kind, both were cases of pyrexia with 1 each after CORBEVAX^TM^ and the XBB.1.5 RBD subunit vaccine. While most AEs were found to be causally related to the test vaccine (58 of 59 AEs) and control vaccine (25 of 27 AEs), the severity was mostly mild (56 of 59 AEs in the test vaccine group and 26 of 27 AEs in the control vaccine group). Adverse event data are presented in a tabular form in Tables 5 and 6 and additional AE data are presented in Supplementary Tables 2 to 5.

**Table 5:** Summary of AEs in the safety population.

| **Parameter/Visit**<br>**N1 (%) (95% CI) n** | **BE's XBB.1.5 Covid-19**<br>**Vaccine (Group 1)**<br>**(N=240)** | **BE's CORBEVAX**<br>**Vaccine (Group 2)**<br>**(N=120)** |
| --- | --- | --- |
| Number (%) of Participants with at least one AE | 54(22.50%)<br>(17.38,28.31),59 | 27(22.50%)<br>(15.38,31.02),27 |
| Number (%) of Participants with at least one AE reported during 60 minutes post-vaccination. | 17(7.08%)<br>(4.18,11.10),18 | 4(3.33%)<br>(0.92,8.31),4 |
| Number (%) of Participants with at least one AE reported during 60 minutes post 1st dose vaccination. | 14(5.83%)<br>(3.23,9.59),14 | 3(2.50%)<br>(0.52,7.13),3 |
| Number (%) of Participants with at least one AE reported during 60 minutes post 2nd dose vaccination. | 4(1.67%)<br>(0.46,4.21),4 | 1(0.83%)<br>(0.02,4.56),1 |
| Number (%) of Participants with at least one AE reported during 7-day diary period. | 47(19.58%)<br>(14.76,25.18),51 | 22(18.33%)<br>(11.86,26.43),22 |
| Number (%) of Participants with at least one AE reported during 7-day diary period post 1st dose of vaccination. | 39(16.25%)<br>(11.82,21.54),40 | 13(10.83%)<br>(5.90,17.81),13 |
| Number (%) of Participants with at least one AE reported during 7-day diary period post 2nd dose of vaccination. | 11(4.58%)<br>(2.31,8.05),11 | 9(7.50%)<br>(3.49,13.76),9 |
| Number (%) of Participants with at least one AE reported during Day 0 to Day 28. | 40(16.67%)<br>(12.18,22.00),41 | 14(11.67%)<br>(6.53,18.80),14 |
| Number (%) of Participants with at least one AE reported during Day 28 to Day 42 | 16(6.67%)<br>(3.86,10.60),16 | 12(10.00%)<br>(5.27,16.82),12 |
| Number (%) of Participants with at least one AE reported during Day 42 to Day 56 | 2(0.83%)<br>(0.10,2.98),2 | 1(0.83%)<br>(0.02,4.56),1 |
| Number (%) of Participants with at least one AE reported during Day 28 to Day 56 | 17(7.08%)<br>(4.18,11.10),18 | 13(10.83%)<br>(5.90,17.81),13 |
| Number (%) of Participants with at least one Solicited AE | 47(19.58%)<br>(14.76,25.18),51 | 22(18.33%)<br>(11.86,26.43),22 |
| Number (%) of Participants with at least one unsolicited AE | 7(2.92%)<br>(1.18,5.92),8 | 5(4.17%)<br>(1.37,9.46),5 |
| Number (%) of Participants with at least one Local AE | 29(12.08%)<br>(8.24,16.89),32 | 17(14.17%)<br>(8.47,21.71),17 |
| Number (%) of Participants with at least one Systemic AE | 26(10.83%)<br>(7.20,15.47),27 | 10(8.33%)<br>(4.07,14.79),10 |
| Number (%) of Participants with at least one Serious AE | 0(0.00%)<br>(0.00,0.00),0 | 0(0.00%)<br>(0.00,0.00),0 |
| Number (%) of Participants with at least one Medically attended AE | 1(0.42%)<br>(0.01,2.30),1 | 1(0.83%)<br>(0.02,4.56),1 |
| Number (%) of Participants with at least one AESI | 0(0.00%)<br>(0.00,0.00),0 | 0(0.00%)<br>(0.00,0.00),0 |
| Number (%) of Participants with Severity grading |  |  |
| Mild | 51(21.25%)<br>(16.25,26.97),56 | 26(21.67%)<br>(14.67,30.11),26 |
| Moderate | 3(1.25%)<br>(0.26,3.61),3 | 1(0.83%)<br>(0.02,4.56),1 |

| Parameter/Visit<br>N1 (%) (95% CI) n | BE's XBB.1.5 Covid-19<br>Vaccine (Group 1)<br>(N=240) | BE's CORBEVAX<br>Vaccine (Group 2)<br>(N=120) |
| --- | --- | --- |
| Severe | 0(0.00%)<br>(0.00,0.00),0 | 0(0.00%)<br>(0.00,0.00),0 |
| Number (%) of Participants with Causality grading |  |  |
| Related | 54(22.50%)<br>(17.38,28.31),58 | 25(20.83%)<br>(13.96,29.20),25 |
| Unrelated | 1(0.42%)<br>(0.01,2.30),1 | 2(1.67%)<br>(0.20,5.89),2 |
| N = No. of participants per arm,<br>N1 = Participant count, % = Percentage of participants, CI = Confidence Interval, n = Event count |  |  |

**Table 6:** Summary of AEs by System-Organ-Class and preferred term in the safety Population.

| SOC/ Preferred Term<br>N1 (%) (95% CI) n | Overall |  |
| --- | --- | --- |
|  | BE's XBB.1.5 Covid-19<br>Vaccine (Group 1) (N=240) | BE's CORBEVAX Vaccine<br>(Group 2) (N=120) |
| Number of Participants with at least one AE | 54(22.50%)<br>(17.38,28.31),59 | 27(22.50%)<br>(15.38,31.02),27 |
| Gastrointestinal disorders | 2(0.83%)<br>(0.10,2.98),2 | 0(0.00%)<br>(0.00,0.00),0 |
| Abdominal Pain Upper | 1(0.42%)<br>(0.01,2.30),1 | 0(0.00%)<br>(0.00,0.00),0 |
| Nausea | 1(0.42%)<br>(0.01,2.30),1 | 0(0.00%)<br>(0.00,0.00),0 |
| General disorders and administration site condition | 43(17.92%)<br>(13.28,23.36),46 | 23(19.17%)<br>(12.56,27.36),23 |
| Fatigue | 1(0.42%)<br>(0.01,2.30),1 | 1(0.83%)<br>(0.02,4.56),1 |
| Injection Site erythema | 2(0.83%)<br>(0.10,2.98),2 | 3(2.50%)<br>(0.52,7.13),3 |
| Injection site Pain | 27(11.25%)<br>(7.55,15.94),30 | 14(11.67%)<br>(6.53,18.80),14 |
| Pyrexia | 13(5.42%)<br>(2.92,9.08),13 | 5(4.17%)<br>(1.37,9.46),5 |
| Nervous System Disorder | 9(3.75%)<br>(1.73,7.00),9 | 3(2.50%)<br>(0.52,7.13),3 |
| Headache | 9(3.75%)<br>(1.73,7.00),9 | 3(2.50%)<br>(0.52,7.13),3 |
| Respiratory, thoracic and mediastinal | 2(0.83%)<br>(0.10,2.98),2 | 0(0.00%)<br>(0.00,0.00),0 |
|  | BE's XBB.1.5 Covid-19<br>Vaccine (Group 1) (N=240) | BE's CORBEVAX Vaccine<br>(Group 2) (N=120) |
| Cough | 1(0.42%)<br>(0.01,2.30),1 | 0(0.00%)<br>(0.00,0.00),0 |
| Rhinorrhoea | 1(0.42%)<br>(0.01,2.30),1 | 0(0.00%)<br>(0.00,0.00),0 |
| Skin and Subcutaneous tissue<br>disorder | 0(0.00%)<br>(0.00,0.00),0 | 1(0.83%)<br>(0.02,4.56),1 |
| Miliaria | 0(0.00%)<br>(0.00,0.00),0 | 1(0.83%)<br>(0.02,4.56),1 |

N = No. of participants per arm, N1 = Participant count, % = Percentage of participants, CI = Confidence Interval, n = Event count

## Discussion

This Phase III study was designed to assess the immunogenicity and safety of a new updated XBB.1.5 subunit vaccine. The test vaccine was compared to Biological E’s CORBEVAX^TM^, a subunit vaccine that has the RBD of the ancestral Wuhan COVID-19 strain. Both these vaccines have been developed on the same platform which uses a *Pichia pastoris* yeast expression system ^16, 17, 18, 19, 20, 21^. The primary objective was to assess if two booster doses of the XBB.1.5 subunit vaccine were superior to two booster doses of the ancestral strain vaccine by XBB.1.5 RBD neutralizing antibody titers and the *a priori* defined superiority criteria were met. Four weeks after the first XBB.1.5 booster, the neutralizing antibody titers were more than twice that in the arm that received the ancestral strain vaccine as a booster. Two weeks after a second booster this differential was nearly three times showing meaningful superiority of the updated XBB.1.5 subunit vaccine over the Ancestral strain RBD subunit vaccine. Secondary immunogenicity endpoints included calculating the GMFR of neutralizing antibodies and anti-XBB.1.5 IgGs as well as the rate of seroconversion by neutralizing antibody titer assessments. More than 89% of the participants that received the updated vaccine demonstrated detectable nAb titers by Day 42, which is two weeks after the second booster. These neutralizing antibodies are of particular importance given that they are likely correlates of protection against COVID-19 infection ^22, 23^. The GMFR of neutralizing antibodies was nearly 8 in the XBB.1.5 arm at Day 28 and more than 17 on Day 42. In comparison, in the ancestral strain the GMFR was about 3 on Day 28 and a little less than 5 on Day 42. Anti-XBB.1.5 IgG GMFRs in the two arms were comparable while being slightly higher after the updated subunit vaccine. Importantly, a *post hoc* analysis showed that participants in this study vaccinated with the XBB.1.5 RBD subunit vaccine also produced neutralizing antibodies against the JN.1 SARS-CoV-2 strain. The test vaccine produced a near two-fold rise in post-boost to pre-boost neutralizing antibodies on Day 42 suggesting robust cross protection against JN.1. These immunogenicity data when taken together support the robust immunogenic response to the XBB.1.5 strain by the updated XBB.1.5 RBD subunit vaccine administered as two booster doses.

Safety-wise, the XBB.1.5 RBD subunit vaccine was found to have a comparable safety profile to CORBEVAX^TM^ which in turn has gone through an extensive clinical development program with nearly 10,000,000 doses having been administered ^11, 12, 13^. The safety profile of the ancestral strain vaccine was found to be consistent with other protein subunit vaccines. Overall, the new XBB.1.5 RBD subunit vaccine was found have an acceptable safety profile when administered in a two dose series across multiple age groups.

Multiple biopharmaceutical companies have developed or are developing monovalent vaccines targeting the XBB.1.5 strain. Presently, the only vaccine targeting the XBB.1.5 strain with Emergency Use Listing from the WHO is NUVAXOVID™ Omicron XBB.1.5 developed by Novavax ^24^. This vaccine, and two other mRNA vaccines from Pfizer and Moderna have received emergency use authorization from the United States FDA ^25^. Pfizer conducted a Phase II/III study to assess the safety and immunogenicity of their XBB.1.5 in participants ≥12 years of age who had received 3 or more doses of an mRNA vaccine with the most recent one being against the BA.4/BA.5-adapted bivalent one. This study showed strong neutralizing antibody response to the Omicron XBB.1.5 strain and the authors concluded that the vaccine had a favourable risk-benefit profile ^26^. Another study showed that after a XBB.1.5 Moderna mRNA booster, the neutralizing antibody titer GMFR against JN.1 was 27 suggesting that vaccines that target XBB.1.5 may also protect against the JN.1 sub-lineage ^27^. A recent preprint supports this notion by showing that nursing home residents in the United States had robust neutralizing antibody GMFRs against both XBB.1.5 and JN.1 strains ^28^ after being vaccinated with a monovalent XBB.1.5 vaccine. Recent observational data also shows that the clinical severity of those vaccinated with a XBB.1.5 COVID-19 vaccine and then infected with the JN.1 strain was not worse than those that acquired the XBB.1.5 strain ^29^. Although this was not a head-to-head study, the GMFR values observed with our vaccine (GMFR of 7.6 at Day 28) are comparable to published data from XBB.1.5 mRNA and protein-based boosters. In a Pfizer Phase II/III clinical trial of a monovalent XBB.1.5 mRNA vaccine, GMFR post vaccination was 6.9 against XBB.1.5 in participants aged between 18 and 55 years of age^24^. Similarly, participants that received Novavax’s protein subunit XBB.1.5 booster had a GMFR of about 7.5 post-vaccination (Lancet Infectious Diseases). In Moderna’s Study 205J, participants that received a monovalent XBB.1.5 mRNA booster as their 5^th^ COVID vaccination had a GMFR of 17.5 (FDA presentation), similar to the Day 42 GMFR of 17 we report where participants had received either 4 or 5 COVID vaccine doses in total.

This Phase III study of a new XBB.1.5 subunit vaccine is limited as it did not include efficacy assessments of the updated booster series. Immunogenicity assessments for this study were conducted in about 360 participants putting the level of evidence generated lower than large efficacy Phase III trials that enroll thousands of participants. While the *a priori* defined superiority objective was met, it remains to be seen if the vaccine reduces the incidence of symptomatic COVID-19 infections, hospitalizations, and deaths due the new strain. Large real-world studies of other XBB.1.5 have shown a reduction in hospitalizations and deaths due to COVID-19 supporting the widespread use of this updated vaccine ^30^. Another important limitation is that participants in this Phase III study were followed up for only 42 days for this study. It is therefore unclear how long these protective neutralizing antibody titers remain for. Our study excluded key risk groups—including individuals with confirmed or suspected immunosuppressive or immunodeficient conditions. These groups are a major focus of COVID-19 vaccination strategies in 2025, which prioritize the prevention of severe disease in immunocompromised individuals. This exclusion limits the generalizability of our findings to certain high-risk populations and warrants explicit mention. A common limitation to all strain-specific booster vaccines and their trials is the ever evolving nature of the SARS-CoV-2 virus. At the time of conduct of this study, JN.1 became the predominant strain in India ^32^. We showed that our vaccine elicited robust cross-protection against the JN.1 strain.

While there is evidence that XBB.1.5-specific vaccines, like ours, also protect against JN.1, it remains to see if these vaccines also provide protection against the later NB.1.8.1 and LF.7 strains. An administrative disadvantage of our XBB.1.5 RBD subunit vaccine was that it tested as a two-dose booster series compared to other single-dose boosters. But, we demonstrate significant neutralizing antibodies post the first booster dose itself. As of writing this manuscript, Biological E is developing a monovalent variant-specific vaccine against the JN.1 strain as recommended by the WHO (SAGE committee recommendation on 23^rd^ December, 2024 and reaffirmed in March 2025). For the clinical development of this vaccine, it will be tested as a single-dose booster likely improving uptake of boosters in general. This aligns with WHO SAGE’s stratified approach to booster vaccination, which prioritises single-dose boosting in high-risk groups—including older adults, individuals with underlying comorbidities, immunocompromised persons, pregnant individuals, and frontline health workers—while permitting flexible, context-dependent use in healthy adults and children. The focus remains on optimising booster coverage in populations at highest risk for severe outcomes, without mandating repeated dosing in low-risk groups. The JN.1 formulation is being pursued based on available immunogenicity and neutralisation data, in line with WHO guidance to select vaccine composition on the basis of prevailing immune escape and variant dominance, particularly in the XBB/JN.1 lineage cluster such as NB.1.8.1 and LF.7.

In conclusion, we provide strong evidence to suggest a favourable risk-benefit profile for the use of Biological E’s updated XBB.1.5 RBD subunit vaccine as a two-dose booster for 5 to 80 year-olds. Based on these data, the vaccine received permission for restricted use in an emergency situation from the Central Drug Standard Control Organization, the Indian national regulatory agency. This vaccine – if approved for use by the WHO – will be particularly important for the rest of the Global South as well, given its lower cost and easier cold-chain management than existing mRNA vaccines. Post-approval real-world and Phase IV studies may provide additional evidence about the continued safety and efficacy of this vaccine.

## Data Availability

All data produced in the present work are contained in the manuscript

## Data Sharing Statement

Access to raw or de-identified participant-level data will be granted to qualified researchers on reasonable request made to the corresponding author and subject to institutional review.

---

**Supplementary table 1:** Demographic and Baseline Characteristics (by age subgroup) - ITT Population.

| Parameter | Statistic | Subgroup 1 |  | Subgroup 2 |  | Subgroup3 |  |
| --- | --- | --- | --- | --- | --- | --- | --- |
|  |  | BE's<br>XBB.1.5<br>Covid-19<br>Vaccine<br>(Group 1)<br>(N=80) | BE's<br>CORBEVAX<br>Vaccine<br>(Group 2)<br>(N=40) | BE's<br>XBB.1.5<br>Covid-19<br>Vaccine<br>(Group 1)<br>(N=80) | BE's<br>CORBEVAX<br>Vaccine<br>(Group 2)<br>(N=40) | BE's<br>XBB.1.5<br>Covid-19<br>Vaccine<br>(Group 1)<br>(N=80) | BE's<br>CORBEVAX<br>Vaccine<br>(Group 2)<br>(N=40) |
| Age<br>(Years) | n | 80 | 40 | 80 | 40 | 80 | 40 |
|  | Mean | 55.4 | 55.8 | 34.1 | 32.5 | 13.0 | 13.4 |
|  | SD | 4.98 | 5.29 | 8.55 | 7.58 | 3.15 | 2.96 |
|  | Median | 53.0 | 54.5 | 33.0 | 32.5 | 14.0 | 14.0 |
|  | IQR (q1:<br>q3) | (52.0,59.0) | (51.0,59.5) | (27.0,40.5) | (26.5,36.0) | (11.0,15.0) | (11.0,16.0) |
|  | Range:<br>min, max | 50.0, 68.0 | 50.0, 68.0 | 19.0, 49.0 | 19.0, 49.0 | 5.0, 17.0 | 6.0, 17.0 |
|  | 95% CI | (54.3,56.5) | (54.1,57.4) | (32.2,36.0) | (30.1,34.9) | (12.3,13.7) | (12.4,14.3) |
| Sex | Male, n<br>(%) | 68(85.0) | 30(75.0) | 63(78.8) | 28(70.0) | 33(41.3) | 20(50.0) |
|  | Female, n<br>(%) | 12(15.0) | 10(25.0) | 17(21.3) | 12(30.0) | 47(58.8) | 20(50.0) |
| BMI (kg/<br>m2) | n | 80 | 40 | 80 | 40 | 80 | 40 |
|  | Mean | 24.9 | 24.7 | 24.4 | 24.4 | 16.3 | 16.7 |
|  | SD | 2.80 | 2.73 | 3.24 | 3.21 | 2.84 | 2.24 |
|  | Median | 25.0 | 24.6 | 25.1 | 23.5 | 16.2 | 16.5 |
|  | IQR (q1:<br>q3) | (23.3,26.8) | (23.0,26.6) | (22.3,26.5) | (21.9,26.5) | (14.5,17.4) | (15.0,18.4) |
|  | Range:<br>min, max | 18.2, 31.7 | 18.8, 29.4 | 16.7, 35.0 | 19.6, 32.5 | 10.4, 27.3 | 12.6, 24.0 |
|  | 95% CI | (24.2,25.5) | (23.8,25.5) | (23.7,25.1) | (23.3,25.4) | (15.6,16.9) | (16.0,17.4) |
| Length<br>(cms) | n | 80 | 40 | 80 | 40 | 80 | 40 |
|  | Mean | 163.9 | 163.5 | 166.2 | 163.8 | 144.7 | 146.4 |
|  | SD | 7.64 | 8.63 | 4.89 | 8.40 | 15.08 | 14.18 |
|  | Median | 166.0 | 165.0 | 167.0 | 164.5 | 148.5 | 149.5 |
|  | IQR (q1:<br>q3) | (160.5,168.5) | (158.0,170.0) | (164.0,170.0) | (158.0,170.0) | (134.0,155.0) | (135.5,157.5) |
|  | Range:<br>min, max | 140.0, 179.0 | 146.0, 183.0 | 147.0, 175.0 | 143.0, 178.0 | 110.0, 170.0 | 115.0, 170.0 |
|  | 95% CI | (162.1,165.6) | (160.7,166.3) | (165.1,167.3) | (161.1,166.5) | (141.3,148.0) | (141.9,150.9) |
| Weight<br>(gms) | n | 80 | 40 | 80 | 40 | 80 | 40 |
|  | Mean | 66.8 | 66.1 | 67.5 | 65.3 | 34.5 | 36.4 |
|  | SD | 8.92 | 10.32 | 9.51 | 9.30 | 9.54 | 9.52 |
|  | Median | 67.1 | 66.6 | 69.2 | 64.3 | 34.6 | 36.0 |
|  |  | BE's XBB.1.5 Covid-19 Vaccine (Group 1) (N=80) | BE's CORBEVAX Vaccine (Group 2) (N=40) | BE's XBB.1.5 Covid-19 Vaccine (Group 1) (N=80) | BE's CORBEVAX Vaccine (Group 2) (N=40) | BE's XBB.1.5 Covid-19 Vaccine (Group 1) (N=80) | BE's CORBEVAX Vaccine (Group 2) (N=40) |
|  | IQR (q1: q3) | (61.1,74.1) | (58.8,72.6) | (61.0,74.6) | (58.7,71.8) | (28.0,40.3) | (30.8,43.2) |
|  | Range: min, max | 42.0, 86.2 | 45.0, 90.4 | 45.0, 84.0 | 45.0, 86.2 | 15.6, 69.0 | 17.5, 60.8 |
|  | 95% CI | (64.8,68.8) | (62.8,69.4) | (65.4,69.6) | (62.3,68.3) | (32.4,36.7) | (33.3,39.4) |
| Ethnicity | Asian, n (%) | 80(100.0) | 40(100.0) | 80(100.0) | 40(100.0) | 80(100.0) | 40(100.0) |

**Supplementary table 2:** Summary of AEs within 60 minutes’ post vaccination by SOC and PT – Safety Population (N=360)

| Parameter<br>N1 (%) (95% CI) n | BE's XBB.1.5 Covid-19 Vaccine<br>(Group 1) (N=240) | BE's CORBEVAX Vaccine<br>(Group 2) (N=120) |
| --- | --- | --- |
| Number of Participants with at least one AE within 60 minutes post vaccination | 17(7.08%)<br>(4.18,11.10),18 | 4(3.33%)<br>(0.92,8.31),4 |
| General Disorders and Administration Site Condition | 17(7.08%)<br>(4.18,11.10),18 | 4(3.33%)<br>(0.92,8.31),4 |
| Injection Site Pain | 17(7.08%)<br>(4.18,11.10),18 | 4(3.33%)<br>(0.92,8.31),4 |

N = No. of participants per arm, N1 = Participant count, % = Percentage of participants, CI = Confidence Interval, n = Event count

**Supplementary table 3:** Summary of Solicited AEs (7-days post vaccination) by SOC and PT – Safety Population (N=360)

| SOC/ Preferred Term<br>N1 (%) (95% CI) n | BE's XBB.1.5 Covid-19 Vaccine (Group 1) (N=240) | BE's CORBEVAX Vaccine (Group 2) (N=120) |
| --- | --- | --- |
| Number of Participants with at least one Solicited AE | 47(19.58%)<br>(14.76,25.18),51 | 22(18.33%)<br>(11.86,26.43),22 |
| General Disorders And Administration Site Condition | 41(17.08%)<br>(12.55,22.45),44 | 20(16.67%)<br>(10.49,24.56),20 |
| Injection Site Erythema | 2(0.83%)<br>(0.10,2.98),2 | 3(2.50%)<br>(0.52,7.13),3 |
| Injection Site Pain | 27(11.25%)<br>(7.55,15.94),30 | 13(10.83%)<br>(5.90,17.81),13 |
| Pyrexia | 12(5.00%)<br>(2.61,8.57),12 | 4(3.33%)<br>(0.92,8.31),4 |

| **SOC/ Preferred Term**<br>**N1 (%) (95% CI) n** | **BE's XBB.1.5 Covid-19**<br>**Vaccine (Group 1)**<br>**(N=240)** | **BE's CORBEVAX**<br>**Vaccine (Group 2)**<br>**(N=120)** |
| --- | --- | --- |
| Nervous System Disorder | 7(2.92%)<br>(1.18,5.92),7 | 2(1.67%)<br>(0.20,5.89),2 |
| Headache | 7(2.92%)<br>(1.18,5.92),7 | 2(1.67%)<br>(0.20,5.89),2 |

N = No. of participants per arm, N1 = Participant count, % = Percentage of participants, CI = Confidence Interval, n = Event count

**Supplementary table 4:** Summary of Unsolicited AEs by SOC and PT – Safety Population (N=360)

| **Parameter**<br>**N1 (%) (95% CI) n** | **BE's XBB.1.5 Covid-19 Vaccine**<br>**(Group 1) (N=240)** | **BE's CORBEVAX Vaccine**<br>**(Group 2) (N=120)** |
| --- | --- | --- |
| Number of Participants with at least one Unsolicited AE | 7(2.92%)<br>(1.18,5.92),8 | 5(4.17%)<br>(1.37,9.46),5 |
| Gastrointestinal Disorders | 2(0.83%)<br>(0.10,2.98),2 | 0(0.00%)<br>(0.00,0.00),0 |
| Abdominal Pain Upper | 1(0.42%)<br>(0.01,2.30),1 | 0(0.00%)<br>(0.00,0.00),0 |
| Nausea | 1(0.42%)<br>(0.01,2.30),1 | 0(0.00%)<br>(0.00,0.00),0 |
| General Disorders And Administration Site Condition | 2(0.83%)<br>(0.10,2.98),2 | 3(2.50%)<br>(0.52,7.13),3 |
| Fatigue | 1(0.42%)<br>(0.01,2.30),1 | 1(0.83%)<br>(0.02,4.56),1 |
| Injection Site Pain | 0(0.00%)<br>(0.00,0.00),0 | 1(0.83%)<br>(0.02,4.56),1 |
| Pyrexia | 1(0.42%)<br>(0.01,2.30),1 | 1(0.83%)<br>(0.02,4.56),1 |
| Nervous System Disorder | 2(0.83%)<br>(0.10,2.98),2 | 1(0.83%)<br>(0.02,4.56),1 |
| Headache | 2(0.83%)<br>(0.10,2.98),2 | 1(0.83%)<br>(0.02,4.56),1 |
| Respiratory, Thoracic And Mediastinal Disorder | 2(0.83%)<br>(0.10,2.98),2 | 0(0.00%)<br>(0.00,0.00),0 |
| Cough | 1(0.42%)<br>(0.01,2.30),1 | 0(0.00%)<br>(0.00,0.00),0 |
| Rhinorrhoea | 1(0.42%)<br>(0.01,2.30),1 | 0(0.00%)<br>(0.00,0.00),0 |
| Skin And Subcutaneous Tissue Disorder | 0(0.00%)<br>(0.00,0.00),0 | 1(0.83%)<br>(0.02,4.56),1 |
| Miliaria | 0(0.00%)<br>(0.00,0.00),0 | 1(0.83%)<br>(0.02,4.56),1 |

N = No. of participants per arm, N1 = Participant count, % = Percentage of participants, CI = Confidence Interval, n = Event count

**Supplementary table 5:** Summary of AEs by SOC and PT, Severity & Causality– Safety Population (N=360)

| Parameter<br>N1 (%) (95% CI) n | Severity | Causality | BE's XBB.1.5 Covid-19<br>Vaccine (Group 1)<br>(N=240) | BE's CORBEVAX<br>Vaccine (Group 2)<br>(N=120) |
| --- | --- | --- | --- | --- |
| Number of Participants<br>with at least one AE |  |  | 54(22.50%)<br>(17.38,28.31),59 | 27(22.50%)<br>(15.38,31.02),27 |
| Gastrointestinal<br>Disorders |  |  | 2(0.83%)<br>(0.10,2.98),2 | 0(0.00%)<br>(0.00,0.00),0 |
| Abdominal Pain Upper |  |  | 1(0.42%)<br>(0.01,2.30),1 | 0(0.00%)<br>(0.00,0.00),0 |
|  | Mild | Possible | 1(0.42%)<br>(0.01,2.30),1 | 0(0.00%)<br>(0.00,0.00),0 |
| Nausea |  |  | 1(0.42%)<br>(0.01,2.30),1 | 0(0.00%)<br>(0.00,0.00),0 |
|  | Mild | Possible | 1(0.42%)<br>(0.01,2.30),1 | 0(0.00%)<br>(0.00,0.00),0 |
| General Disorders And<br>Administration Site<br>Condition |  |  | 43(17.92%)<br>(13.28,23.36),46 | 23(19.17%)<br>(12.56,27.36),23 |
| Fatigue |  |  | 1(0.42%)<br>(0.01,2.30),1 | 1(0.83%)<br>(0.02,4.56),1 |
|  | Mild | Possible | 1(0.42%)<br>(0.01,2.30),1 | 0(0.00%)<br>(0.00,0.00),0 |
|  | Mild | Probable | 0(0.00%)<br>(0.00,0.00),0 | 1(0.83%)<br>(0.02,4.56),1 |
| Injection Site Erythema |  |  | 2(0.83%)<br>(0.10,2.98),2 | 3(2.50%)<br>(0.52,7.13),3 |
|  | Mild | Probable | 2(0.83%)<br>(0.10,2.98),2 | 3(2.50%)<br>(0.52,7.13),3 |
| Injection Site Pain |  |  | 27(11.25%)<br>(7.55,15.94),30 | 14(11.67%)<br>(6.53,18.80),14 |
|  | Mild | Certain | 17(7.08%)<br>(4.18,11.10),18 | 4(3.33%)<br>(0.92,8.31),4 |
|  | Mild | Possible | 7(2.92%)<br>(1.18,5.92),9 | 8(6.67%)<br>(2.92,12.71),8 |
|  | Mild | Probable | 3(1.25%)<br>(0.26,3.61),3 | 2(1.67%)<br>(0.20,5.89),2 |
| Pyrexia |  |  | 13(5.42%)<br>(2.92,9.08),13 | 5(4.17%)<br>(1.37,9.46),5 |
|  | Mild | Certain | 3(1.25%)(0.26,3.61),3 | 0(0.00%)<br>(0.00,0.00),0 |
|  | Mild | Possible | 1(0.42%)(0.01,2.30),1 | 1(0.83%)(0.02,4.56),1 |
|  | Mild | Probable | 6(2.50%)(0.92,5.36),6 | 3(2.50%)(0.52,7.13),3 |
|  | Mild | Unlikely | 0(0.0%) | 1(0.83%)(0.02,4.56),1 |
|  | Moderate | Possible | 1(0.42%)(0.01,2.30),1 | 0(0.0%) |
|  | Moderate | Probable | 2(0.83%)(0.10,2.98),2 | 0(0.0%) |
| Nervous System<br>Disorder |  |  | 9(3.75%)<br>(1.73,7.00),9 | 3(2.50%)<br>(0.52,7.13),3 |
| Headache |  |  | 9(3.75%)<br>(1.73,7.00),9 | 3(2.50%)<br>(0.52,7.13),3 |
|  | Mild | Certain | 3(1.25%)<br>(0.26,3.61),3 | 1(0.83%)<br>(0.02,4.56),1 |
|  | Mild | Possible | 3(1.25%)<br>(0.26,3.61),3 | 1(0.83%)<br>(0.02,4.56),1 |
|  | Mild | Probable | 3(1.25%)<br>(0.26,3.61),3 | 0(0.00%)<br>(0.00,0.00),0 |
|  | Moderate | Probable | 0(0.00%)<br>(0.00,0.00),0 | 1(0.83%)<br>(0.02,4.56),1 |
| Respiratory, Thoracic<br>And Mediastinal<br>Disorder |  |  | 2(0.83%)<br>(0.10,2.98),2 | 0(0.00%)<br>(0.00,0.00),0 |
| Cough |  |  | 1(0.42%)<br>(0.01,2.30),1 | 0(0.00%)<br>(0.00,0.00),0 |
|  | Mild | Possible | 1(0.42%)<br>(0.01,2.30),1 | 0(0.00%)<br>(0.00,0.00),0 |
| Rhinorrhoea |  |  | 1(0.42%)<br>(0.01,2.30),1 | 0(0.00%)<br>(0.00,0.00),0 |
|  | Mild | Unlikely | 1(0.42%)<br>(0.01,2.30),1 | 0(0.00%)<br>(0.00,0.00),0 |
| Skin And Subcutaneous<br>Tissue Disorder |  |  | 0(0.00%)<br>(0.00,0.00),0 | 1(0.83%)<br>(0.02,4.56),1 |
| Miliaria |  |  | 0(0.00%)<br>(0.00,0.00),0 | 1(0.83%)<br>(0.02,4.56),1 |
|  | Mild | Unrelated | 0(0.00%)<br>(0.00,0.00),0 | 1(0.83%)<br>(0.02,4.56),1 |

N = No. of participants per arm, N1 = Participant count, % = Percentage of participants, CI = Confidence Interval, n = Event count

**Supplementary table 6:** Summary of MAAEs by SOC and PT – Safety Population (N=360)

| Parameter<br>N1 (%) (95% CI) n | BE's XBB.1.5 Covid-19<br>Vaccine (Group 1) (N=240) | BE's CORBEVAX Vaccine<br>(Group 2) (N=120) |
| --- | --- | --- |
| Number of Participants with at least<br>one MAAE | 1(0.42%)<br>(0.01,2.30),1 | 1(0.83%)<br>(0.02,4.56),1 |
| General Disorders And Administration<br>Site Condition | 1(0.42%)<br>(0.01,2.30),1 | 1(0.83%)<br>(0.02,4.56),1 |
| Pyrexia | 1(0.42%)<br>(0.01,2.30),1 | 1(0.83%)<br>(0.02,4.56),1 |

N = No. of participants per arm, N1 = Participant count, % = Percentage of participants, CI = Confidence Interval, n = Event count

